# H_2_ inhalation therapy in patients with moderate Covid 19 (H_2_ COVID): a prospective ascending-dose phase 1 clinical trial

**DOI:** 10.1101/2024.03.15.24304071

**Authors:** Cordélia Salomez-Ihl, Joris Giai, Maud Barbado, Adeline Paris, Saber Touati, Jean- Pierre Alcaraz, Stéphane Tanguy, Corentin Leroy, Audrey Lehmann, Bruno Degano, Marylaure Gavard, Pierrick Bedouch, Patricia Pavese, Alexandre Moreau-Gaudry, Mathieu Roustit, François Boucher, Philippe Cinquin, Jean-Paul Brion

## Abstract

**Introduction:** The Covid-19 pandemic, caused by the Severe Acute Respiratory Syndrome Coronavirus 2 (SARS-CoV-2), has triggered a serious global health crisis, resulting in millions of reported deaths since its initial identification in China in November 2019. The global disparities in immunization access emphasize the urgent need for ongoing research into therapeutic interventions. This study focuses on the potential use of molecular dihydrogen (H2) inhalation as an adjunctive treatment for Covid-19. H2 therapy shows promise in inhibiting intracellular signaling pathways associated with inflammation, particularly when administered early in conjunction with nasal oxygen therapy.

**Methods:** This Phase I study, characterized by an open-label, prospective, monocentric, and single ascending dose design, seeks to assess the safety and tolerability of the procedure in individuals with confirmed SARS-CoV-2 infection. Employing a 3+3 design, the study includes three exposure durations (target durations): 1 day (D1), 3 days (D2), and 6 days (D3).

**Results:** We concluded that the Maximum Tolerated Duration is at least three days. Every patient showed clinical improvement and excellent tolerance to H2 therapy.

**Discussion/conclusion:** To the best of our knowledge, this phase 1 clinical trial is the first to establish the safety of inhaling a mixture of H2 (3.6%) and N2 (96.4%) in hospitalized Covid-19 patients. The original device and method employed ensure the absence of explosion risk. The encouraging outcomes observed in the 12 patients included in the study justify further exploration through larger, controlled clinical trials.

**Question:** What is the Maximum Tolerated Duration of inhalation for a gaseous mixture including 3.6% of molecular dihydrogen in moderate COVID-19 patients?

**Findings:** The Maximum Tolerated Duration is at least three days. All patients showed clinical improvement and excellent tolerance to H2 therapy. To the best of our knowledge, this phase 1 clinical trial is the first to establish the safety of inhaling a mixture of H2 (3.6%) and N2 (96.4%) in hospitalized Covid-19 patients.

**Meaning:** A gaseous mixture including 3.6% H2, considered in the literature to have promising anti-inflammatory potential, and presenting no risk of explosion, can be used in patients with moderate COVID 19 for at least three days.

## INTRODUCTION

Severe acute respiratory syndrome coronavirus 2 (SARS-CoV-2) has been responsible for coronavirus disease 2019 (Covid-19) since November 2019 when it was first discovered in China. Since then, more than 770 million cases and almost 7 million deaths have been reported around the world [1]. This pathology comes with life-threatening respiratory symptoms in severe cases, especially in patients with risk factors such as age, obesity, diabetes, and cardiovascular diseases [2]. Since the appearance of this pathology, epidemiological data have evolved thanks to the development of vaccination. Indeed, the vaccine protects against severe forms and has contributed to a significant reduction in hospitalizations and deaths [3]. Nevertheless, access to immunization shows significant disparities across the world [4]. Therefore, it remains essential to continue the research effort for therapeutic strategies.

In this context, dihydrogen (H_2_) inhalation could be an interesting opportunity. Hyperbaric H_2_ inhalation was first described in the 1970s [5] to have potential for cancer treatment, and the first preclinical study at atmospheric pressure dates back to 2007 [6], in a model of cerebral infarction in the rat. Since then, Ito et al. [7] have shown that H_2_ inhibits intracellular signaling pathways of inflammation without involving anti-free radical effects. In addition, H_2_inhalation (2.9%) has also been reported to limit mast cell activation [8]. Xie et al. have shown that two 60 min sessions of inhalation of a gas mixture containing 2% H_2_ allow limitation of multiple organ damage and mortality in a model of generalized inflammation in mice [9]. They also have shown that inhaling H_2_ restores the PaO_2_ / FiO_2_ ratio, both in a mouse model of sepsis by cecal ligation [10] and in a model of lung damage induced by lipopolysaccharides [11]. H_2_ has also been described as reducing the significant burden on lung parenchyma during Covid-19 [12]. In view of the current data in the literature, the application of a H_2_ treatment makes it possible to trigger numerous potentially protective mechanisms in a hyperinflammatory context, such as sepsis and very probably Covid-19, by trapping hydroxyl radicals and peroxynitrite, by limiting inflammatory reactions by modulating intracellular transduction cascades and by modifying the expression of certain genes [13].

About tolerance, H_2_ has been safely used in the sixties at very high doses, to prevent decompression sickness and arterial gas thrombi, in deep-diving gas mixes (Hydreliox = breathing gas mixture used at high pressure – 60 bars- containing 49% H_2_, 50% helium and 1% O_2_) [14]. In the clinical context, H_2_ has been shown to have no effect on temperature, blood pressure, pH or SpO_2_ [15]. In humans, no adverse effects related to H_2_ have been described with H_2_ inhalation in hundreds of patients until now [16].

Concerning administration, several routes have been considered. The most widely used today, both in pre-clinical and clinical trials, are the ingestion of hydrogen-enriched drinking water and the inhalation of a gas mixtures [17]. H_2_ is considered highly flammable when its concentration in the air exceeds 4.1% [18]. As a result, until recently, the used gas mixtures all contained between 2% and 4% H_2_.

In Spring 2020, when we initiated this research, anti-inflammatory strategies such as corticosteroids were the only drugs that showed efficacy in Covid-19 patients. Chinese guidelines already recommended use of H_2_ in Covid-19 patient management [19,20]. Then, a Chinese team published results of use of H_2_ in 2020 in an efficacy open label clinical trial with the administration of a mixture including 67% H_2_ and 33% of O_2_, with statistically significant improvement of clinical and biological parameters [21]. Since then, other studies using inhalation of mixtures with 66% of H_2_ in acute and post-acute Covid-19 have been launched [22,23]. However, the explosion hazard is not discussed, and these mixtures do not comply with norms and regulations in several countries. Our hypothesis is that the administration of H_2_ mixtures below the explosivity level of 4.1% could safely improve the clinical condition of hospitalized moderate Covid-19 patients (WHO clinical progression scale 5 [24]). The primary aim of this study was to establish the feasibility and safety of an original protocol of H_2_ inhalation, by defining its Maximum Tolerated Duration (MTD).

## METHODOLOGY

### Study design

This Phase I, open-label, prospective, monocentric, single ascending dose study aims to establish the safety and the tolerability of the procedure in patients with confirmed SARS-Cov-2 infection. A 3+3 design was used, with 3 durations of exposure (target durations): 1 day (D1), 3 days (D2) and 6 days (D3), as summarized in Figure 1 and in Supplemental Data. This study was approved by French National Agency for Drug Safety (ANSM) and has been approved by a personal protection committee (clinicaltrials.gov identifier NCT04633980).

**Figure 1.**
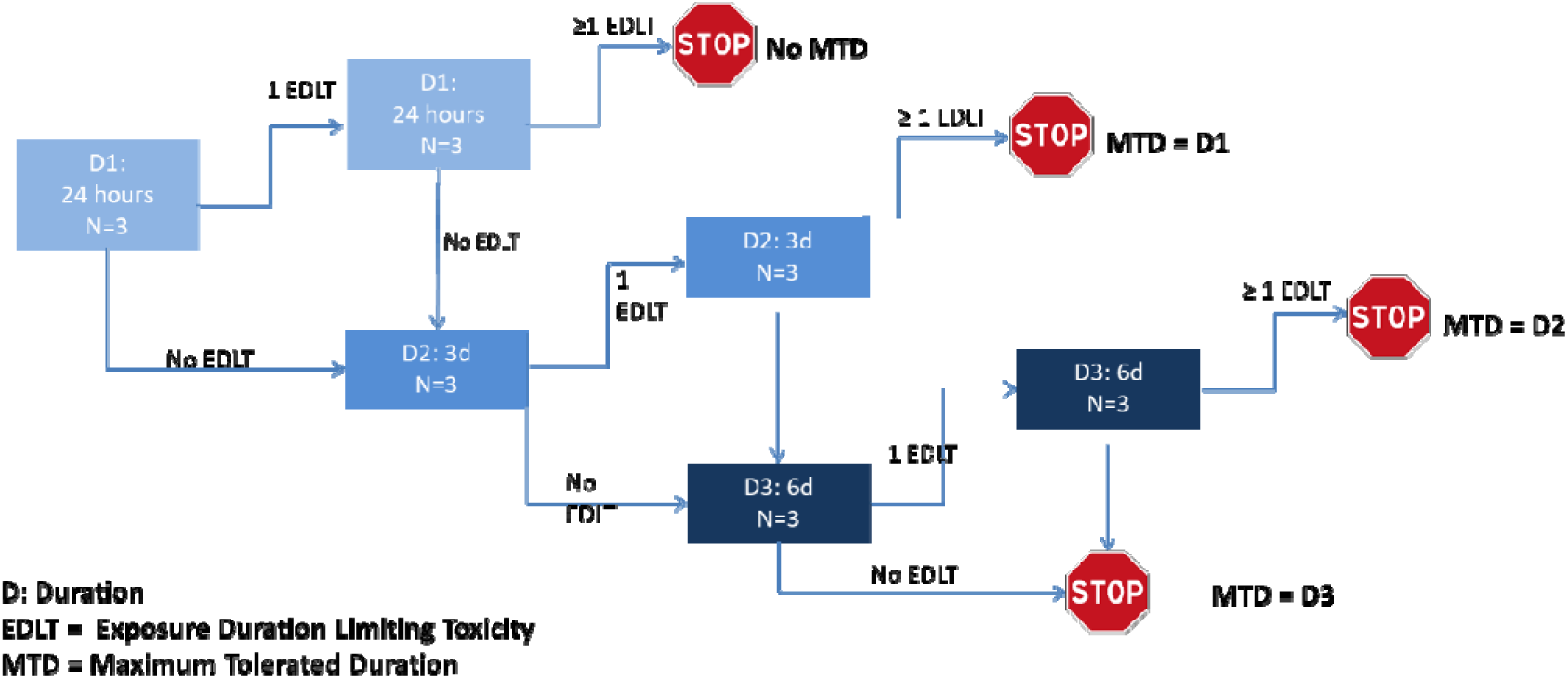
Design of the single ascending dose study conducted in patients, with 3 doses (target durations) tested (D1 to D3).

### Study population

We included adult patients with suspicion of SARS CoV-2 infection based on clinical signs and positive PCR, and hospitalized with SpO_2_ ≤94% on room air requiring normobaric oxygen therapy with a nasal flow of O_2_ ≤ 6L /min to reach at least SaO_2_ ≥ 95%. Detailed inclusion and exclusion criteria are specified in Supplemental Data.

### Interventions

All patients received the usual standard of care during their hospitalization (antibiotics, systemic corticosteroid therapy and preventive anticoagulation).

An original medical delivery device has been designed by our team and has undergone a risk analysis by an independent organization. This device includes a flow regulator (a CE-marked medical device for clinical trials) allowing to guarantee a fixed flow of 1 L/min of a specific medical grade gas mixture (3.6% H_2_; 96.4% N_2_), manufactured and supplied by AIR PRODUCTS, packaged in B50 type cylinders. The gas mixture is combined with O_2_ from the oxygen outlet of the wall (O_2_ flow adapted to the needs of the patient in accordance to standard of care). The device and method ensure that there is no risk of explosion or ignition. The medical delivery device is illustrated in Figure 2. If a clinical and radiological improvement occurred, and if SpO_2_ remained above 95%, O_2_ and H_2_ inhalation were stopped even if the target duration of 24 hours, 3 or 6 days was not achieved.

**Figure 2.**
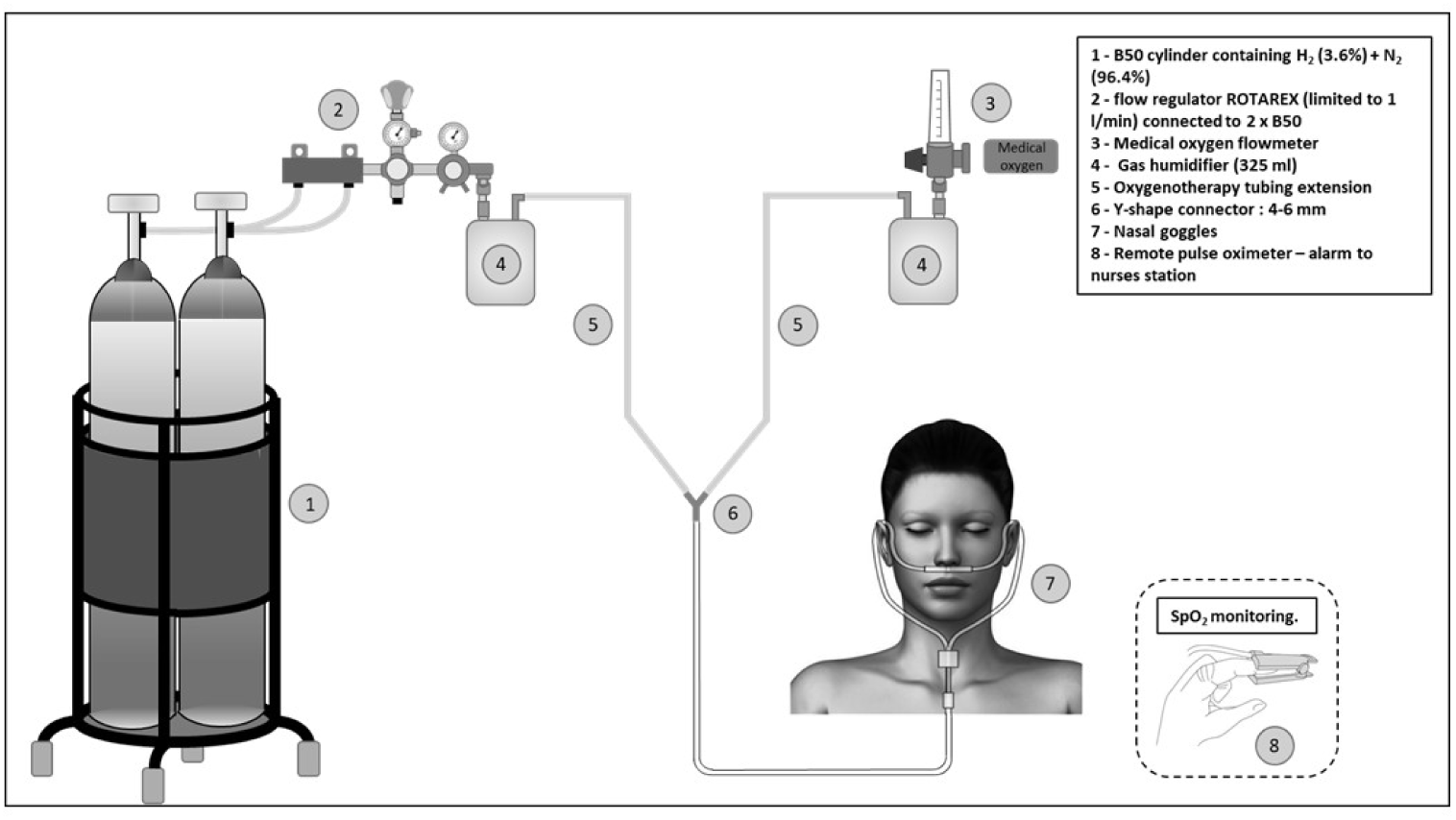
Overview of Hydrogen Inhalation in Covid patients

Specific training has been given to user personnel, and information has been provided to guarantee suitability for use (posters, explanatory documents).

### Outcomes

Because the concentration and the flow of the inhaled mixture is kept constant, the term “dose” actually refers to the target duration of exposure to H_2_ inhalation. The classical notion of dose-limiting toxicity (DLT) therefore becomes “Exposure Duration – Limiting Toxicity” (EDLT), and is defined as the occurrence of any of the following Serious Adverse Event (SAE) rated according to the NIH Common Terminology Criteria for Adverse Events (CTCAE 5.0) [25] during and over 3 days after the end of H_2_:

- observed grade >=4 toxicity from the “Respiratory, thoracic and mediastinal disorders” section of CTCAE v5.0;
- OR observed grade >=3 toxicity from other sections of CTCAE v5.0;
- OR any relevant deterioration in the health of the subject;
-AND at least possibly related with H_2_.

The primary outcome of this study is the MTD, defined as the maximum duration of exposure to H_2_ with no more than one EDLT. If an EDLT occurs, three additional patients with not EDLT have to be included to authorize moving to the next step. As a consequence, between 6 and 24 patients could have been included in the study (see Table 1 and Figure 1).

**Table 1.** Population description.

| **Cohort** | **1 day treatment**<br>N = 3 | **3 days treatment**<br>N = 3 | **6 days treatment**<br>N = 6 | **Total**<br>N = 12 |
| --- | --- | --- | --- | --- |
| Age |  |  |  |  |
| Mean (SD) | 52.67 ( 6.66) | 56.33 ( 1.53) | 62.33 (14.11) | 58.42 ( 10.57) |
| Median | 56.00 | 56.00 | 64.50 | 57.50 |
| Q1-Q3 | 50.50 - 56.50 | 55.50 - 57.00 | 57.00 - 72.75 | 55.75 - 61.25 |
| Min-Max | 45.00 - 57.00 | 55.00 - 58.00 | 39.00 - 76.00 | 39.00 - 76.00 |
| N | 3 | 3 | 6 |  |
| Sex |  |  |  |  |
| Female | 1 (33.3%) | 0 ( 0.0%) | 0 ( 0.0%) | 1 ( 8.3%) |
| Male | 2 (66.7%) | 3 (100.0%) | 6 (100.0%) | 11 (91.7%) |
| (col %) | N = 3 | N = 3 | N = 6 | N = 12 |
| weight (kg) |  |  |  |  |
| Mean (SD) | 88.00 ( 8.19) | 77.33 ( 11.15) | 77.50 ( 16.13) | 80.08 ( 13.81) |
| Median | 90.00 | 73.00 | 77.50 | 81.25 |
| Q1-Q3 | 84.50 - 92.50 | 71.00 - 81.50 | 63.75 - 86.75 | 71.25 - 90.00 |
| Min-Max | 79.00 - 95.00 | 69.00 - 90.00 | 60.00 - 101.00 | 60.00 - 101.00 |
| N | 3 | 3 | 6 |  |
| Height (cm) |  |  |  |  |
| Mean (SD) | 173.00 ( 11.36) | 177.33 ( 3.21) | 176.50 ( 5.96) | 175.83 ( 6.83) |
| Median | 178.00 | 176.00 | 177.50 | 177.50 |
| Q1-Q3 | 169.00 - 179.50 | 175.50 - 178.50 | 172.50 - 181.00 | 174.00 - 181.00 |
| Min-Max | 160.00 - 181.00 | 175.00 - 181.00 | 168.00 - 183.00 | 160.00 - 183.00 |
| N | 3 | 3 | 6 |  |
| BMI (kg/m2) |  |  |  |  |
| Mean (SD) | 29.67 ( 5.03) | 24.47 ( 2.66) | 24.68 ( 3.89) | 25.88 ( 4.84) |
| Median | 29.00 | 23.80 | 23.75 | 24.50 |
| Q1-Q3 | 27.00 - 32.00 | 23.00 - 25.60 | 21.88 - 27.20 | 22.58 - 28.50 |
| Min-Max | 25.00 - 35.00 | 22.20 - 27.40 | 20.50 - 30.50 | 20.50 - 35.00 |
| N | 3 | 3 | 6 |  |

An independent clinical events committee (CEC) was constituted: i) to review all adverse events before each duration increase; ii) each time a SAE occurred, in order to assess the imputability of H_2_; iii) on investigator or sponsor demand. The study had to be discontinued anytime on CEC request, particularly if a SAE were attributed to the intervention, which would have immediately stopped the study.

The secondary outcomes consist in the analysis of serum levels of CRP, lymphocytes, and lymphocytes/thrombocytes.

### Statistical analysis

The study population was the intention to treat population, i.e. patients were analyzed in the initial cohort they were allocated to, even if H_2_ therapy was stopped prematurely. Descriptive statistics were performed for all evaluation criteria. Categorical variables were presented using counts and frequencies, while continuous variables were presented using mean, standard deviation, minimum, median, maximum, interquartile range, and number of subjects with evaluable data. Normality of continuous variables was assessed graphically. Analysis was performed with R software (version ≥ 4.2).

## RESULTS

### Description of population

Twelve participants were recruited from the Department of Infectious Diseases at the University Hospital of Grenoble, from 19^th^ January 2021 to 31^st^ May 2022. The last patient follow-up was 7*^th^*June 2022. Three received one day of treatment and three received three days of treatment. Six patients were recruited for the 6-day treatment. Among them two were treated for the entire planned duration and four prematurely discontinued the treatment because their clinical improvement was such that they no longer required oxygen-therapy and were discharged from the hospital.

Overall, eleven patients were male (92%), the mean age was 58 (*SD =10.8*) and the mean body mass index was 26 (*SD =4.2*).

A CT scan (iodin injection was done if pulmonary embolism was suspected) was performed at inclusion for all patients except two. In all cases, it showed specific images of Covid-19 pneumonia and the proportion of damage was found between 25 and 50%.

On average, patients were included 12.2 days after the onset of their symptoms (SD*= 2.0*) [minimum: 9 days, maximum: 16 days].

### Primary outcome

The Maximum Tolerated Duration was at least three days, since only two out of the 6 patients included in step 3 of the study (D3, 6-day treatment) were treated for 6 days. Indeed, the clinical condition of the other four patients included improved so well before D3 that discontinuation of oxygen therapy was decided before the end of this period, so that this step could not be validated. Two SAE occurred. The first one in a patient whose oxygen requirement increased during the 3-day observation period after the end of H_2_ inhalation. This patient was admitted during 24h in intensive care unit, received up to 40L/min of O_2_ supplement, and rapidly recovered. The second SAE occurred in a patient with a pulmonary embolism, which did not preclude the continuation of the H_2_ therapy. Both SAE were attributed to Covid-19, and not to H_2_, by the CEC.

### Secondary outcomes

#### Effect of H_2_ treatment on clinical status (Chair Rise test)

The evaluation of the physical condition of the patients was based on the Chair Rise test, due to its specificity. 12 patients started the chair rise test, but only 6 completed the test (the other tests were stopped because of patient SpO_2_ desaturation). For the patients who completed the test, the initial moderate desaturation (SpO2 = 95%) decreased to 92% at the end of the test.

All 12 patients were able to perform this test at the end of the treatment + 3 days. Four patients still needed O_2_ supplementation and showed desaturation on the complete test. The mean flow of O_2_ for these 4 patients was 3 ± 0.1 L/min.

#### Effect of H_2_ treatment on biological variables: CRP evolution is illustrated in Figure 3

Lymphocyte evolution is illustrated in Figure 4.

**Figure 3.**
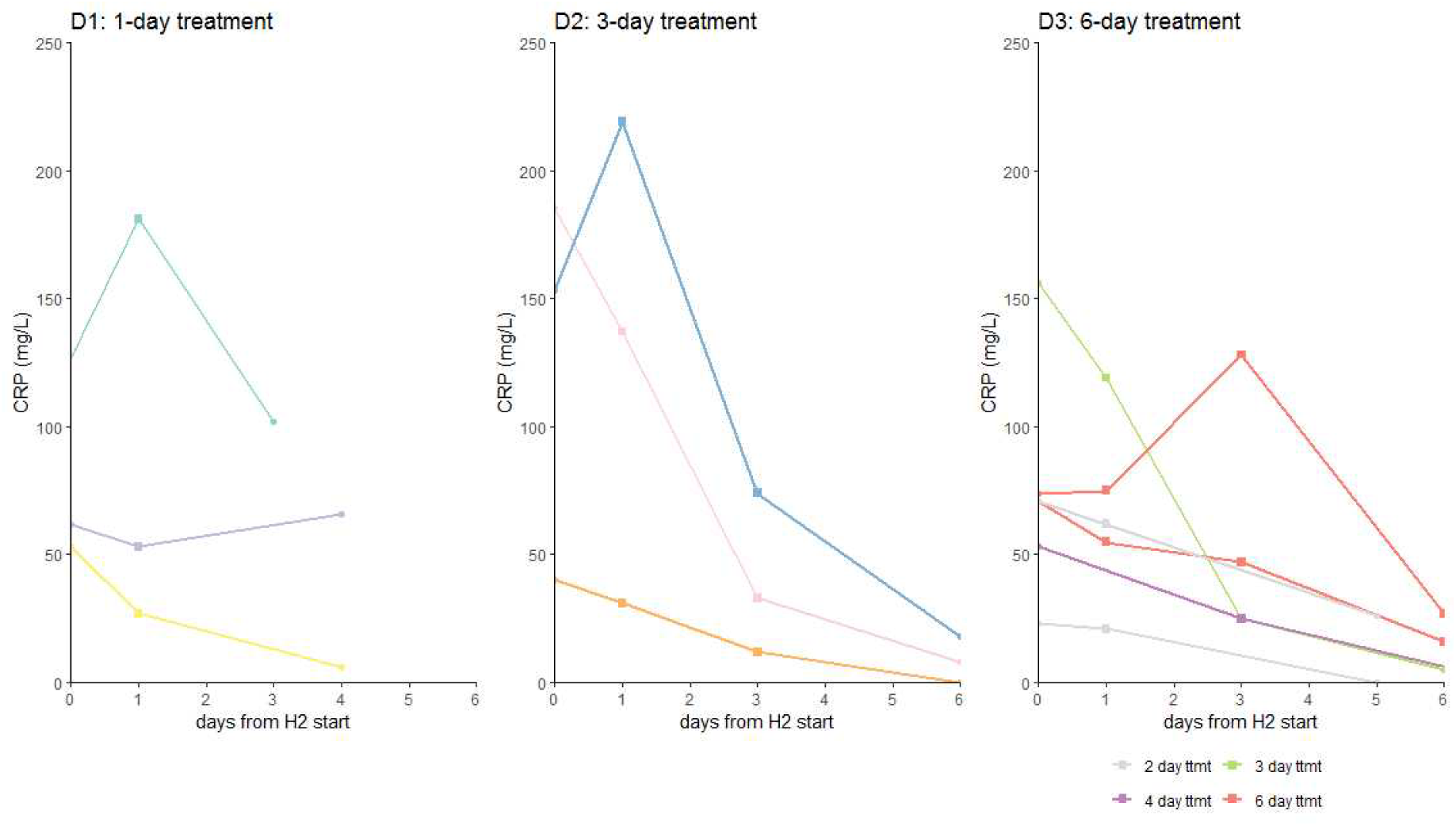
CRP evolution (mg/L) according to the number of days of treatment

**Figure 4.**
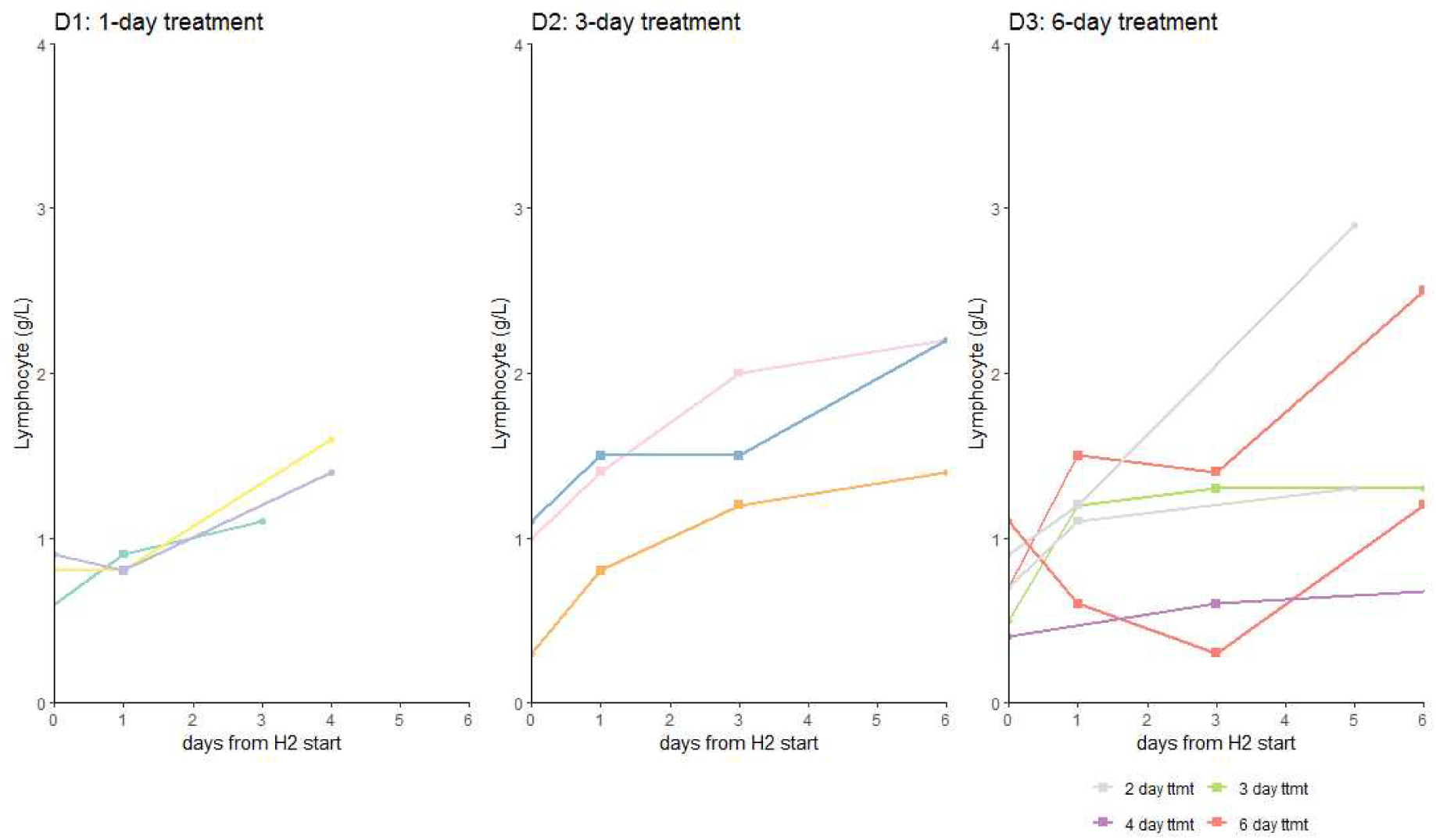
Lymphocyte evolution (G/L) according to the number of days of treatment

## DISCUSSION

All patients who improved clinically tolerated H_2_ therapy perfectly. This study is, to our knowledge, the first phase 1 clinical trial to demonstrate the safety of the inhalation of a H_2_ (3.6%) - N_2_ (96.4%) mixture in hospitalized Covid-19 patients, with an original device and method guaranteeing the absence of explosion risk. We demonstrated a MTD of at least three days, which seems to be sufficient for Covid-19 WHO-scale 5 patient management, since clinical improvement is significant at day 3.

Our results remain very encouraging concerning longer durations of treatment, since none of the six patients treated in the D3 group of our study presented any adverse effects attributed to H_2_. All observed adverse events are complications of moderate Covid-19, well described in the literature [26, 27].

Concerning secondary outcomes, the methodology of a phase one test does not allow us to conclude on the possible efficacy of the gas mixture. However, we have very encouraging results.

Figure 3 shows a trend towards a decrease in CRP. The mean decrease in CRP at day 3 was 18 for D1 patients, 86 for D2 patients, and 28 for D3 patients. This contrasts with the literature, which reports a stagnation in the first days after hospitalization of patients at stage 5 of the WHO scale [28]. Of course, due to the limited number of patients, these results would require confirmation by a phase 2 study.

Lymphopenia is usually more common in patients with COVID-19 [29]. For example, Huang et al. evaluated the data of 36 patients who died due to COVID-19 in 2020. 70.59% had lymphopenia [30]. In our study, on the contrary, patients seem to have increased their lymphocytes during gas-mixture therapy.

The clinical and biological observations of our study are consistent with the literature, considering that H_2_ has been described as able to reduce lung injury and thus to reduce the number of critically ill patients [31]. Another literature review has explained that H_2_ could directly enter the lung tissue through ventilatory activities and exert anti-inflammatory effects at the multiple stages of the inflammatory response, alleviating the airway damage caused by the excessive activation of the inflammatory cells and the massive release of inflammatory factors [32]. In addition, during Covid-19 associated pulmonary injury, activation of resident alveolar macrophages has led to the release of potent proinflammatory mediators and chemokines that promote the accumulation of neutrophils and monocytes [33]. Inhaled H_2_ exerts a non-specific anti-inflammatory effect on macrophages, neutrophils and lymphocytes and inhibits ROS production [34].

The designed delivery device guaranteed a fixed flow rate of 1 L/min of the gas mixture, while allowing the adaptation of the O_2_ flow rate to the patient’s needs. Since this flow is completed by patient’s natural breathing, there is no risk of suffocation of the patient, even if the device is improperly used. At a concentration of 3.6% of H_2_ in the mixture, the patient receives 1.5 mmol/min 24 hours a day, corresponding to 2160 mmol/day. This dose is significantly lower than the dose administered in the protocols where the patient inhales a stoichiometric mixture H_2_ 66% - O_2_ 33%. However, the question of the explosion hazard associated to the clinical use of this stoichiometric mixture is not addressed in the corresponding publications. Literature data [15] have led us to suggest that much lower concentrations, respecting the safety standards accepted in the majority of countries, have an anti-inflammatory activity and therefore could have an efficacy against Covid-19 comparable to that reported in China.

Finally, demonstrating the safety of an H_2_ inhalation protocol compatible with explosion risk standards opens up the possibility of ambulatory use of H_2_ gas. Beyond Covid-19, there is a considerable potential for the combined ambulatory administration of O_2_ and H_2_ to the lungs. Indeed, H_2_ is the only known molecule with anti-inflammatory properties that is totally devoid of recognized adverse effects.

## CONCLUSION

We demonstrated that H_2_ inhalation at 3.6% delivered with our device is a safe therapy in humans, including those with viral pulmonary pathology. This clinical trial is the first step towards approval of our H_2_ inhalation protocol as a drug delivered by a medical device. More data are obviously awaited. In particular, it would be important to carry out phase 2 and 3 clinical trials, with a much larger number of patients, in order to demonstrate H_2_ efficacy in the management of pathologies involving oxidative and inflammatory phenomena, including of course COVID19. Pathologies with strong pulmonary inflammatory component, such as Chronic Obstructive Pulmonary Disease, could also benefit greatly from this potential therapy.

## Data Availability

All data produced in the present study are available upon reasonable request to the authors.

